# Targeted Muscle Reinnervation (TMR) or Regenerative Peripheral Nerve Interface (RPNI) for pain prevention in patients with limb amputation: a protocol for a systematic review and meta-analysis

**DOI:** 10.1101/2024.12.23.24319539

**Authors:** Jesús del Moral Preciado, David Gurpegui, Montserrat Royo, Bernardo Hontanilla

## Abstract

**Introduction:** Regenerative Peripheral Nerve Interface (RPNI) and Targeted Muscle Reinnervation (TMR) are two reinnervation techniques which have shown clear superiority over classical amputation. It is mainly due to a lower incidence of painful neuromas, residual limb pain and phantom limb pain associated with these new procedures.

However, they have never been compared to each other. Neither has their effectiveness been evaluated based on patients demographics, age, sex, comorbidities (diabetes, coronary heart disease, peripheral arterial disease, chronic kidney disease, congestive heart failure), amputations cause, type of amputation, amputation level, previous surgeries and if there was or not previous nerve division into fascicles. Therefore, the objective of this systematic review and meta-analysis is to compile all the evidence to date and provide a comprehensive view of what each technique offers.

**Methods and design:** The review will be conducted according to this protocol, following the recommendations of the ‘Cochrane Handbook for Systematic Reviews’. A comprehensive electronic search will be performed in: Cochrane Register of Controlled Trials (CENTRAL), Web of Science, Scopus, PubMed and MedRixb. This review will include randomized, quasi-randomized, and observational studies written in any language. We will use Covidence for assessing all titles and abstracts identified during the literature search. Two review authors will independently assess the trial eligibility, risk of bias and extract appropriate data points.

**Ethics and dissemination:** The proposed systematic review will collect and analyse data from published studies; therefore, it raises no ethical issues. The results of the review will be disseminated by publication in a peer-review journal and submitted for presentations at conferences.

**PROSPERO registration number:** CRD42024617299

**STRENGTHS AND LIMITATIONS OF THIS STUDY:**

- This will be the first systematic review to include a comparison between RPNI and TMR.
- Through a comprehensive search and selection of high-quality articles, the best available evidence of RPNI and TMR against classical amputation will be gathered.
- Gray literature and unpublished studies will be sourced from MedRixb aiming to reduce the impact of a possible publication bias.
- Exclusion of non-English/Spanish papers may lead to language bias.

**PICO QUESTION:** *Population:* Any adults (aged over 18 years) and gender with a superior or inferior limb amputation.

*Intervention:* Prophylactic Targeted Muscle Reinnervation (TMR) or Regenerative Peripheral Nerve Interface (RPNI).

*Comparators:* Classical amputation. TMR vs RPNI.

*Outcomes:* 1) Incidence of neuroma, 2) Incidence of residual limb pain (RLP), 3) Severity of Pain 4) Incidence of phantom limb pain (PLP), and 5) Surgical complications (dehiscence, infection, haematoma and seroma).

## INTRODUCTION

### Description of the condition

Limb amputation has an estimated incidence of 185,000 persons each year in the United States (1) with a tendency towards increasing prevalence (2). This significant increase is widely accepted to be caused by the ageing of the population and an increasing incidence of diabetes mellitus and vascular disease (2).

Amputations still cause significant morbidity and pain (3). Amputations are associated with depression as a reaction to the sudden disability (4), surgical site infection, increased risk of thromboembolism in patients subjected to a major amputation (5–8). And a significant risk of perioperative morbidity and mortality, generally in patients with vascular disease (9). In fact, the overall 5-year mortality rate in patients with any amputation ranged from 53% to 100% and from 52% to 80% for patients with major amputations, being age, renal disease, proximal amputation, diabetes, and peripheral vascular disease the risk factors. This indicates amputation as a possible prognostic severity marker (10).

Some demographic factors have shown a higher risk of incidence of amputations, Black and Latino patients being up to four (11), and one and a half (12) times respectively more likely to receive amputations when compared to white patients.

The most common aetiologies for lower limb loss are: 1) vascular pathology (mainly ischaemia due to peripheral artery disease or embolism, and diabetes mellitus), 2) trauma, 3) cancer, and 4) congenital abnormalities. On the other hand, aetiologies for upper extremity amputation in adults are 1) trauma, 2) ischaemia, 3) infection, and 4) malignancy (2,13,14).

Nerves are completely encased in an epineural sheath, which is made of well-organized fibrous connective tissue. Significant disruption of this sheath occurs during (the process of) an amputation, causing disorganized regeneration of the enclosed axons, and proliferation into scar and connective tissue, resulting in an amputation neuroma (15,16). This type of neuroma belongs to a class of tumours referred to as reactive tumours, which are considered as benign non-neoplastic nerve tumours. Although benign, they may be symptomatic, evolving into severe pathology, generating pain with numbness or paresthesias, decreasing quality of life, forcing medication and preventing prosthetic use (15–18).

Though benign, they can become symptomatic, transforming into severe pathologies by producing

Some authors have described a series of criteria to classify neuromas as symptomatic (16). See **Table 1**.

**Table 1.** Arnold’s criteria for classifying neuromas as symptomatic.

| **Table 1.** Arnold's criteria for classifying neuromas as symptomatic |
| --- |
| **Mandatory criteria (must be all 3):**<br>1. Pain with at least 3 of the following characteristics: burning, sharp, shooting, electric, paresthesias, numbness, cold intolerance.<br>2. Symptoms in a defined neural anatomic distribution.<br>3. History of nerve injury or suspected nerve injury. |
| **Partial criteria (at least 1):**<br>1. Positive Tinel Sign (for cutaneous nerve).<br>2. Positive response to local anaesthetic injection.<br>3. US or MRI confirmation of neuroma. |

Although the incidence of symptomatic neuromas is not fully known, multiple studies have indicated that it is in a range between 4% and 49%. This varies depending on the affected limb, the height of the amputation, previous comorbidities, cause of surgery, and the patient’s follow-up time. (16,18–23).

The pain associated with limb amputation is known as residual limb pain (RLP) or phantom limb pain (PLP). The mechanisms and pathophysiology of RLP and PLP are different and have not been fully elucidated. Peripheral mechanisms include spontaneous activity in dorsal root ganglion neurons and ectopic neural activity originating from afferent fibres in a neuroma (24,25), while central mechanisms include spinal cord sensitization and cortical reorganization (26–29). Both residual limb pain and phantom limb pain can occur independently or concurrently in the post-operative period. It is thought that RLP may result from neuromas in the stump and PLP may be more closely related to cortical reorganization (13,30).

PLP is defined as the perception of sensations, including pain, in an amputated limb or a part of the body that is no longer present. PLP is a common sequel of amputation, occurring in up to 60 to 80% of people with limb amputation in the early post-operative period, with the incidence decreasing with time following amputation. PLP incidence appears to be independent of age, sex, amputation level or side. Prevention by peripheral analgesia has not yielded consistent results (26,31–33).

RLP is defined as a type of pain felt in the part of a limb that remains after an amputation. Acute residual limb pain is a strong predictor of chronic residual limb pain, while age, sex, amputation level, aetiology and postsurgical pain medication quantity do not correlate with chronic residual limb pain (30,34,35).

The psychological impact of these two entities should not be overlooked. People with limb amputations usually experience increased anxiety, depressive symptoms, post-traumatic psychological stress symptoms and affective distress (36,37).

De Lange et al. (38) and Mauch et al. (39) demonstrated that prophylactically intervention with TMR or RPNI improved both residual limb pain and phantom limb pain post-amputation. It appears that performing these techniques does not increase the complication rate beyond prolonging surgical time. However, the superiority of one of the two techniques remains to be demonstrated, as studies comparing them are still scarce. We aim to group all these studies to estimate whether one technique is superior to the other in pain prevention or presents a lower complication rate.

### Description of the interventions

TMR is increasingly being used to treat symptomatic neuromas. In this procedure, severed motor nerves are transferred to the motor branches of nearby healthy muscle segments, which, after reinnervation, gain the capacity of contraction in response to neural control signals (40– 42). Due to the creation of multiple additional control sites, the contraction of target muscles and the functional control of the prosthesis become easier and more intuitive. This way, it has been demonstrated that TMR presents clear benefits compared to classical amputation, although its ability to reduce the incidence of neuromas, as well as the treatment of PLP and RLP, is still being investigated (41–43).

Other authors, on the other hand, carry out this technique by rerouting the severed nerve endings to nearby expendable motor nerve branches of functionally expendable muscles, providing a pathway for axonal growth to these nerves. This serves as biological amplifiers of neural information and limits the regeneration that leads to neuroma formation (44,45).

RPNI involves taking a free muscle graft from the amputated limb (either proximal or distal to the site of amputation) and encasing the muscle graft around the dissected nerve, allowing for greater control over nerve growth to motor end plates on the grafted tissue ((46–59)).

Those autologous free muscle grafts are designed to be small enough to have an easy revascularization and to provide living motor end plates suitable for reinnervation in approximately 1–3 months. It has a great potential for permitting the use of advanced neuroprosthetics or sensory feedback (46,60–64).

TMR is more demanding regarding identification and dissection, as well as needing a prolonged operative time. Some authors believe the morbidity of an autologous muscle graft harvest is lesser than that of the compromise of a motor branch in the residual limb. Furthermore, the risk of a mismatch in nerve calibres is a potential source for axonal escape, which may result in a symptomatic neuroma or into a loss of reinnervation (64–68). In both procedures, the nerve can reinnervate and remodel itself within a denervated muscle segment, which provides many targets for reinnervation and reduces the likelihood of disorganized axonal sprouting, subsequently decreasing the risk of symptomatic neuroma formation (15,41,46,52).

It is important to mention that there have not been studies or systematic reviews which compare TMR vs RPNI, so these are only technical aspects, not evidence of superiority.

### Objectives

Primary objectives

- To determine the efficacy and safety of TMR and RPNI in treating patients with limb amputation.
- To compare which technique (TMR and RPNI) shows better results of efficacy and safety in treating patients with limb amputations.

Secondary objectives

- To determine the efficacy and safety of TMR and RPNI across multiple relevant patient characteristics. These include demography of the patients, age, sex, comorbidities, amputation cause(s), amputation level, previous surgeries and if there was or not previous nerve division into fascicles.

## METHODS

For this protocol, we have followed methodological guidance from the Cochrane Handbook for Systematic Reviews of Interventions (69) and we used the PRISMA-P checklist when writing our report (70,71). For the review, we will follow methodological guidance from the Cochrane Handbook for Systematic Reviews of Interventions (69) and MECIR (Methodological Expectations for Cochrane Intervention Reviews) (72), and report the review following PRISMA (73,74).

### Criteria for considering studies for this review

#### Studies

This review will include all studies written in any language, randomized and non-randomized, which involves clinical trials, cohort studies, case control studies, quasi-experimental studies and case series (with n>10).

We will exclude case reports (detailed reports of an individual or handful of individuals), case series (with n<10), qualitative studies (studies that collect and analyse non-numerical data) and narrative reviews.

#### Participants

We will consider for inclusion participants that are adults (aged over 18 years) of any gender, and with a superior or inferior major limb amputation of any aetiology.

#### Interventions

We will include the following five comparisons.

- Targeted Muscle Reinnervation (TMR) versus classical amputation without further nerve considerations.
- Regenerative Peripheral Nerve Interface (RPNI) against classical amputation without further nerve considerations.
- Targeted Muscle Reinnervation versus regenerative Peripheral Nerve Interface.
- Prophylactic versus curative TMR/RPNI
- Case series (n>10) of prophylactic TMR/RPNI

#### Outcome measures

As no core outcome set has been established for effectiveness trials in amputation reinnervation, a list of outcomes was generated based on existing systematic reviews and primary observational studies identified through a preliminary search of the literature (De Lange et al. (38), Mauch et al. (39), Zimbulis et al. (75), and Yuan et al. (76)). The primary outcomes are neuroma incidence (present or not present), pain, phantom limb pain (PLP) and residual limb pain (RLP) (with PROMIS and VAS scales, and if it is or not present). All surgical complications will be analysed as secondary outcomes (dehiscence, infection, haematoma, seroma, abscess, cardiac events, wound healing complications, acute kidney disease, superficial wounds, operative stump revisions and last mortality rate).

### Search methods for identification of studies

We will search the following electronic databases:

- Web of Science (inception to present).
- Scopus (inception to present).
- PubMed (inception to present).
- Cochrane Register of Controlled Trials (CENTRAL) (inception to present).
- CINAHL (inception to present).
- Google Scholar (inception to present).

To identify additional published, unpublished, and ongoing studies, we will:

- Check the reference lists of all relevant studies.
- Search in MedRxiv (inception to present)
- Contact authors, if appropriate.

Online supplemental material 1 demonstrates the complete search strategy.

## DATA COLLECTION AND ANALYSIS

### Selection of studies

We will assess all titles and abstracts identified during the literature searches, using Covidence (77). Two review authors (JM, MR) will independently review the search results and identify reports by screening all remaining titles and abstracts, excluding those that do not meet inclusion criteria. Afterwards, we will retrieve the full text of the articles, and both review authors will independently screen them for relevance and extract data. We will reach out to the investigators to retrieve missing information when necessary. Any disagreements will be resolved by discussion with the third author (DG).

The reasons for exclusion will be documented and presented at the full-text stage under the title of ‘Characteristics of excluded studies’.

### Data extraction and management

The following data will be extracted from each study

- Administrative details: Study author(s), date of publication, year in which study was conducted and presence of conflict of interest.
- Study characteristics: Study registration, study design type and sample size.
- Participants: Age, demography, sex, number of patients randomized, number lost to follow-up/withdrawn, number analysed, previous surgeries and comorbidities.
- Interventions: Type of amputation, cause of amputation, level of amputation and if there was or not previous nerve division into fascicles.
- Outcomes: Incidence of neuroma, incidence of PLP, incidence of RLP, severity of pain and incidence of surgical complications.

Analysis will be performed with STATA 16 metan command (78).

### Risk of bias assessment in included studies

Screening, data extraction and assessment of the risk of bias of included studies will be done using validated tools and discussed below. Disagreements will be settled by consensus with the fourth author (BH). Risk of bias will be estimated using recommendations in the Cochrane Handbook for Systematic Reviews of Interventions, Version 6.5. Assessment of the risk of bias in randomized trials will be estimated with the Risk of Bias 2 (RoB 2) tool (79). Assessment of the risk of bias in a non-randomized study will be estimated with the Risk Of Bias In Non-randomized Studies of Interventions version 2 (ROBINS-I V2) tool (80). Assessment of the risk of bias in case series studies will be estimated with The Joanna Briggs Institute Checklist for Case Series (81).

### Measures of treatment effect

Measures of treatment effect have been chosen following the recommendations of Cochrane Handbook for Systematic Reviews of Interventions, Version 6.5 (69). Dichotomous outcomes will be synthesized and reported as an odds ratio (OR) with 95% confidence intervals (CI). Continuous outcome data will be measured using mean difference with 95% confidence intervals.

### Dealing with missing data

The researchers of the original studies will be contacted to request any missing data, such as sample sizes for analysis and actual numbers for outcomes presented in the figures. If this proves to be unfeasible, the data will be excluded.

### Assessment of heterogeneity and data analysis

The data will be firstly assessed for heterogeneity by visual appraisal of the presented forest plots, and the I^2^ index will be used to quantitatively evaluate heterogeneity among included studies in each forest plot (82). Afterward, a meta-analysis will be performed with DerSimonian– Laird method and random effects model (83).

If sufficient studies are available, we will undertake the following subgroup analyses as follows:

- Sex.
- Demography (ethnicity).
- Cause of amputation is of great relevance as it is associated with the patient’s risk of mortality, the severity of the situation, other complications, etc.
- Type of amputation.
- Amputation level.
- Presence of comorbidities (diabetes, coronary heart disease, peripheral arterial disease, chronic kidney disease or congestive heart failure).
- Previous nerve division into fascicles (it reduces the formation of aberrant reinnervation and promotes proper reinnervation).

A cumulative meta-analysis will also be performed based on the year in which each study was conducted (84).

For studying the age of participants, a meta-regression will be performed.

### Assessment of reporting/publication biases

We will conduct a systematic, comprehensive search of the literature for both published and unpublished studies to minimize the risk of publication bias.

If 10 or more studies are available. We will explore reporting bias graphically using a funnel plot, and statistically by using the Egger’s test to assess the symmetry of the funnel plot. Trim and fill analysis will be performed to further assess the possible publication bias.

### Sensitivity analysis

We will conduct sensitivity analyses to explore the methodological quality of studies, check whether studies with a high risk of bias (in at least two domains) overestimate the effect of treatment and to assess the robustness of our findings.

We will also perform a sensitivity analysis using the data from case series studies and the prophylactic arms of studies comparing prophylactic vs. curative TMR/RPNI, to enhance the sample size.

### Summary of findings and assessment of the certainty of the evidence

We will use the GRADE approach, as outlined in the GRADE Handbook, to assess the certainty of evidence regarding the clinical irrelevant outcomes outlined earlier in this document (85).

### Ethics and dissemination

No ethical approval is needed for this review, as we will use data from previously published studies. We will spread our research in national and international medical congresses and any other non-conflicting means of publication.

## Data Availability

This is just the protocol, we will make our data available when we collect it.

## Acknowledgments

The authors would like to thank Dr. Miguel Ruiz Canela (Professor of Preventive Medicine and Public Health, University of Navarra) for his assistance advising us in the methods section.

## FOOTNOTES

### Author statement

DG is the guarantor of the review, JM and DG initiated the review, drafted and finalised the background, objectives and methods sections. All authors read and approved the final version of the manuscript.

### Patient consent for publication

Not required.

