## Supplementary material for "Targeted Muscle Reinnervation (TMR) or Regenerative Peripheral Nerve Interface (RPNI) for pain prevention in patients with limb amputation: a protocol for a systematic review and meta-analysis": Search strategy (from another version of this preprint: version 1, 2024-12-26)

**Pubmed**

#1 Targeted muscle reinnervation [Title/Abstract]

#2 Targeted muscle re-innervation [Title/Abstract]

#3 TMR [Title/Abstract]

#4 Regenerative peripheral nerve interface [Title/Abstract]

#5 RPNI [Title/Abstract]

#6 Limb amputation [Title/Abstract]

#7 Extremity amputation [Title/Abstract]

#8 Amputation [Title/Abstract]

#9 Amputee [Title/Abstract]

#10 Amputate [Title/Abstract]

#11 Amputation, surgical [MeSH Terms]

#12 Limb disarticulation [Title/Abstract]

#13 Extremity disarticulation [Title/Abstract]

#14 Disarticulation [Title/Abstract]

#15 #1 OR #2 OR #3

#16 #4 OR #5

#17 #6 OR #7 OR #8 OR #9 OR #10 OR #11 OR #12 OR #13 OR #14

#18 #15 AND #16

#19 (#15 OR #16) AND #17

#18

**(((Targeted muscle reinnervation[Title/Abstract]) OR (Targeted muscle re-innervation[Title/Abstract])) OR (TMR[Title/Abstract])) AND ((Regenerative peripheral nerve interface[Title/Abstract]) OR (RPNI[Title/Abstract]))**

**11/12/2024	56 results**

#19

**((((Targeted muscle reinnervation[Title/Abstract]) OR (Targeted muscle re-innervation[Title/Abstract])) OR (TMR[Title/Abstract])) OR ((Regenerative peripheral nerve interface[Title/Abstract]) OR (RPNI[Title/Abstract]))) AND (((((((((Limb amputation[Title/Abstract]) OR (Extremity amputation[Title/Abstract])) OR (Amputation[Title/Abstract])) OR (Amputee[Title/Abstract])) OR (Amputate[Title/Abstract])) OR (Amputation, surgical[MeSH Terms])) OR (Limb disarticulation[Title/Abstract])) OR (Extremity disarticulation[Title/Abstract])) OR (Disarticulation[Title/Abstract]))**

**11/12/2024	268 results**

**Cochrane Register of Controlled Trials (CENTRAL)**

((("Targeted muscle reinnervation":ti,ab) OR ("Targeted muscle re-innervation":ti,ab)) OR (TMR:ti,ab)) AND (("Regenerative peripheral nerve interface":ti,ab) OR (RPNI:ti,ab))

**11/12/2024	6 Trials matching**

(((("Targeted muscle reinnervation":ti,ab) OR ("Targeted muscle re-innervation":ti,ab)) OR (TMR:ti,ab)) OR (("Regenerative peripheral nerve interface":ti,ab) OR (RPNI:ti,ab))) AND ((((((((("Limb amputation":ti,ab) OR ("Extremity amputation":ti,ab)) OR (Amputation:ti,ab)) OR (Amputee:ti,ab)) OR (Amputate:ti,ab)) OR ([mh "Amputation, surgical"])) OR ("Limb disarticulation":ti,ab)) OR ("Extremity disarticulation":ti,ab)) OR (Disarticulation:ti,ab))

**11/12/2024	0 Results**

**Web of Science**

((((TI="Targeted muscle reinnervation" OR AB="Targeted muscle reinnervation")) OR ((TI="Targeted muscle re-innervation" OR AB="Targeted muscle re-innervation"))) OR ((TI=TMR OR AB=TMR))) AND (((TI="Regenerative peripheral nerve interface" OR AB="Regenerative peripheral nerve interface")) OR ((TI=RPNI OR AB=RPNI)))

**11/12/2024	53 Results**

**20/12/2024	54 Results**

(((((TI="Targeted muscle reinnervation" OR AB="Targeted muscle reinnervation")) OR ((TI="Targeted muscle re-innervation" OR AB="Targeted muscle re-innervation"))) OR ((TI=TMR OR AB=TMR))) OR (((TI="Regenerative peripheral nerve interface" OR AB="Regenerative peripheral nerve interface")) OR ((TI=RPNI OR AB=RPNI)))) AND ((((((((((TI="Limb amputation" OR AB="Limb amputation")) OR ((TI="Extremity amputation" OR AB="Extremity amputation"))) OR ((TI=Amputation OR AB=Amputation))) OR ((TI=Amputee OR AB=Amputee))) OR ((TI=Amputate OR AB=Amputate))) OR (ALL="Amputation, surgical")) OR ((TI="Limb disarticulation" OR AB="Limb disarticulation"))) OR ((TI="Extremity disarticulation" OR AB="Extremity disarticulation"))) OR ((TI=Disarticulation OR AB=Disarticulation)))

**11/12/2024	306 Results**

**20/12/2024	307 Results**

**Scopus**

(((TITLE-ABS("Targeted muscle reinnervation")) OR (TITLE-ABS("Targeted muscle re-innervation"))) OR (TITLE-ABS(TMR))) AND ((TITLE-ABS("Regenerative peripheral nerve interface")) OR (TITLE-ABS(RPNI)))

**11/12/2024	66 documents found**

((((TITLE-ABS("Targeted muscle reinnervation")) OR (TITLE-ABS("Targeted muscle re-innervation"))) OR (TITLE-ABS(TMR))) OR ((TITLE-ABS("Regenerative peripheral nerve interface")) OR (TITLE-ABS(RPNI)))) AND (((((((((TITLE-ABS("Limb amputation")) OR (TITLE-ABS("Extremity amputation"))) OR (TITLE-ABS(Amputation))) OR (TITLE-ABS(Amputee))) OR (TITLE-ABS(Amputate))) OR (INDEXTERMS("Amputation, surgical"))) OR (TITLE-ABS("Limb disarticulation"))) OR (TITLE-ABS("Extremity disarticulation"))) OR (TITLE-ABS(Disarticulation)))

**11/12/2024	351 documents found**

**20/12/2024	352 documents found**

**MedRixv**

Abstract or title "Targeted muscle reinnervation" (match all words)

**1 Result 11/12/2024**

Abstract or title "Targeted muscle re-innervation" (match all words)

**No Results 11/12/2024**

Abstract or title "TMR" (match all words)

**13 Results 11/12/2024**

Abstract or title "Regenerative peripheral nerve interface" (match all words)

**No Results 11/12/2024**

Abstract or title "RPNI" (match all words)

**No Results 11/12/2024**
